# Metabolomic and Inflammatory Signatures of Symptom Dimensions in Major Depression

**DOI:** 10.1101/2021.08.05.21261388

**Authors:** Christopher R Brydges, Sudeepa Bhattacharyya, Siamak Mahmoudian Dehkordi, Yuri Milaneschi, Brenda Penninx, Rick Jansen, Bruce S. Kristal, Xianlin Han, Matthias Arnold, Gabi Kastenmüller, Mandakh Bekhbat, Helen S Mayberg, W Edward Craighead, A John Rush, Oliver Fiehn, Boadie W Dunlop, Rima Kaddurah-Daouk, Mood Disorders Precision Medicine Consortium

## Abstract

**Background:** Major depressive disorder (MDD) is a highly heterogenous disease, both in terms of clinical profiles and pathobiological alterations. Recently, immunometabolic dysregulations were shown to be correlated with atypical, energy-related symptoms but less so with the Melancholic or Anxious distress symptom dimensions of depression in The Netherlands Study of Depression and Anxiety (NESDA) study. In this study, we aimed to replicate these immunometabolic associations and to characterize the metabolomic correlates of each of the three MDD dimensions.

**Methods:** Using three clinical rating scales, Melancholic, and Anxious distress, and Immunometabolic (IMD) dimensions were characterized in 158 patients who participated in the Predictors of Remission to Individual and Combined Treatments (PReDICT) study and from whom plasma and serum samples were available. The NESDA-defined inflammatory index, a composite measure of interleukin-6 and C-reactive protein, was measured from pre-treatment plasma samples and a metabolomic profile was defined using serum samples analyzed on three metabolomics platforms targeting fatty acids and complex lipids, amino acids, acylcarnitines, and gut microbiome-derived metabolites among other metabolites of central metabolism.

**Results:** The IMD clinical dimension and the inflammatory index were positively correlated (r=0.19, p=.019) after controlling for age, sex, and body mass index, whereas the Melancholic and Anxious distress dimensions were not, replicating the previous NESDA findings. The three symptom dimensions had distinct metabolomic signatures using both univariate and set enrichment statistics. IMD severity correlated mainly with gut-derived metabolites and a few acylcarnitines and long chain saturated free fatty acids. Melancholia severity was significantly correlated with several phosphatidylcholines, primarily the ether-linked variety, lysophosphatidylcholines, as well as several amino acids. Anxious distress severity correlated with several medium and long chain free fatty acids, both saturated and polyunsaturated ones, sphingomyelins, as well as several amino acids and bile acids.

**Conclusion:** The IMD dimension of depression is reliably associated with markers of inflammation. Metabolomics provides powerful tools to inform about depression heterogeneity and molecular mechanisms related to clinical dimensions in MDD, which include a link to gut microbiome and lipids implicated in membrane structure and function.

## INTRODUCTION

Major depressive disorder (MDD) is an important public health problem, with a lifetime incidence of 16.6% (Kessler et al., 2005). Although MDD has been defined by a consistent set of clinical criteria since the introduction of the DSM-III in 1980, the problem of clinical heterogeneity across individuals has severely hampered progress in achieving a more precise, biologically-based classification system for the syndrome (Fried and Nesse 2015, Park, Kim et al. 2017, Zimmerman, Martin et al. 2019). This clinical heterogeneity likely arises from substantial biological variability which, if better understood, could yield more personalized treatments for patients presenting with the MDD syndrome (Chen, Eaton et al. 2000, Goldberg 2011, Milaneschi, Lamers et al.2016).

Blood-based metabolomic analyses, which measure the presence of small molecules (e.g., amino acids, lipids, organic acids) in the systemic circulation, body fluids and tissues, is a powerful emerging tool for parsing heterogeneous complex disorders. Previous work by our group and others have identified correlations between serum/plasma metabolomic profiles and clinically defined features of MDD including response to treatment (Kaddurah-Daouk, Bogdanov et al. 2013, Bhattacharyya, Ahmed et al. 2019, Ahmed, MahmoudianDehkordi et al. 2020, MahmoudianDehkordi, Ahmed et al. 2021). Prior work has focused primarily on metabolomic analyses conducted on a single analytic platform (Brydges et al., in submission). Analysis of samples across multiple metabolomic platforms that assess a broad range of chemical reactions including gut-derived metabolites offers the promise of a more thorough biological characterization of distinct symptom dimensions that comprise MDD.

The importance of inflammation on MDD symptoms has recently gained increased attention. Epidemiologic data indicate MDD patients are at increased risk of inflammation-related comorbidities, including obesity and metabolic syndrome, cardiovascular disease, and diabetes mellitus (Otte, Gold et al. 2016). These elevated risks are not fully explained by lifestyle factors, such as smoking and a sedentary lifestyle (Penninx 2017). Many MDD patients demonstrate increased circulating levels of pro-inflammatory proteins, including interleukin (IL)-6, and C-reactive protein (CRP) (Dowlati, Herrmann et al. 2010, Kiecolt-Glaser, Derry et al. 2015, Kohler-Forsberg, Buttenschon et al. 2017, Haroon, Daguanno et al. 2018). Clinical trials suggest that the presence of high inflammation in MDD impedes response to monoamine modulating antidepressants and such patients may benefit from more specific anti-inflammatory treatments (Raison, Rutherford et al. 2013, Liao, Xie et al. 2019).

Two historically important clinical dimensions in MDD are melancholic features and anxiety. The clinical features of Melancholia, characterized by psychomotor changes, guilt, and the neurovegetative symptoms of early insomnia plus appetite or weight loss, have long been recognized as an important clinical subtype of depression (Kendler 2020). Anxious distress is characterized by prominent worry or dread, feeling tense, and restlessness (Zimmerman, Martin et al. 2019). Multiple studies have identified poorer treatment outcomes among patients with anxiety (Fava, Rush et al. 2006, Howland, Rush et al. 2009, Gaspersz, Lamers et al. 2017) and mixed results in patients with melancholia (McGrath et al., 2008; Gili et al., 2012; Imai et al., 2021), suggesting that more efficacious treatments will require a deeper understanding of the biological characteristics of these MDD dimensions.

Recently, in the Netherlands Study of Depression and Anxiety (NESDA) cohort of depressed adults, an immunometabolic dimension (IMD) was identified, consisting of the atypical and energy related symptoms of increased appetite, weight gain, hypersomnia, low energy, and leaden paralysis (Lamers, Milaneschi et al. 2018, Alshehri, Boone et al. 2019). Severity of IMD symptoms in this cohort associated with a measure of inflammation derived from serum CRP and IL-6 concentrations (Lamers, Milaneschi et al. 2018, Lamers, Milaneschi et al. 2019, Lamers, Milaneschi et al. 2020, Milaneschi, Lamers et al. 2020). Importantly, the association of inflammation with the IMD dimension was specific, with no significant association found with the Melancholia or Anxious distress dimensions (Lamers, Milaneschi et al. 2020). It should be noted that the crux of IMD is both atypical energy-related symptoms *and* biological inflammatory metabolic dysregulation, as opposed to just the symptomology. Although others have also found associations between related symptom profiles and inflammation (Simmons, Burrows et al. 2020), there have been no attempts to replicate the IMD specific findings of the NESDA analyses.

In the current study, we assessed both inflammatory and metabolomic markers as they relate to the three MDD dimensions of IMD, Melancholia, and Anxious distress. Using samples from a randomized controlled trial of treatment-naïve adults with MDD (Dunlop, Binder et al. 2012), we aimed to: 1) replicate the association between immune-inflammatory dysregulation and IMD severity and the non-association between immune-inflammatory dysregulation and severity of Melancholia and Anxious distress symptoms reported by the NESDA group (Lamers, Milaneschi et al. 2020); and 2) characterize the metabolomic signature of IMD, Melancholia, and Anxious distress symptoms of depression.

## Methods

### Study Design

The Predictors of Remission in Depression to Individual and Combined Treatments (PReDICT) study was conducted from 2007-2013 through the Mood and Anxiety Disorders Program of Emory University. The study was approved by the Institutional Review Board at Emory University and all patients provided written informed consent before participating. The PReDICT study protocol (Dunlop, Binder et al. 2012) and clinical results (Dunlop, Kelley et al. 2017) have been published previously.

Briefly, PReDICT randomly assigned treatment-naïve adult outpatients with non-psychotic MDD to one of three treatments: duloxetine (30-60 mg/day), escitalopram (10-20 mg/day), or CBT (16 one-hour individual sessions) for 12 weeks. Eligibility required a score ≥ 18 at screening and ≥ 15 at baseline on the 17-item Hamilton Depression Rating Scale (HAM-D) (Hamilton 1960). Participants were excluded if they had any medically significant or unstable medication condition that could impact study participation, safety, or data interpretation, any current eating disorder, or any current substance abuse or dependence.

### Symptom Assessments

At the baseline and post-treatment visits, patients were assessed by trained interviewers using the HAM-D 24-item with atypical symptoms, the Hamilton Anxiety Rating Scale (HAMA) (Hamilton 1959), and the Quick Inventory of Depressive Symptomatology Self-Report (QIDS-SR) (Rush, Trivedi et al. 2003). The NESDA study derived the three dimensions of MDD evaluated here based on items from the Inventory of Depressive Symptoms Self-Report (IDS-SR) (Rush et al., 1996) and Beck Anxiety Inventory (Beck et al., 1988). The IDS-SR, QIDS-SR, and BAI items are all scored from 0-3. In order to replicate the NESDA analyses, to construct the A/ER and Melancholia dimensions in PReDICT we used the same items from the QIDS-SR where possible and substituted similar items from the HAMD as described below.

For the IMD symptom dimension, the NESDA study summed the five IDS item scores for hypersomnia, increased appetite, increased weight, low energy, and leaden paralysis (Lamers, Milaneschi et al. 2020). The first four of these symptoms are present in the QIDS-SR used in PReDICT (items 4, 7, 9, and 14, respectively). For the IDS-SR leaden paralysis item, we substituted item E (fatiguability) of the HAM-D. This last item was recoded such that scores of 3 and 4 were replaced with values of 2 and 3, respectively, so that each item was on the same scale and to maintain the overall clinical IMD symptom score range of 0-15. A higher score indicated more severe IMD symptomology.

The Melancholia score was defined in NESDA by summing the 8 IDS-SR items for early morning insomnia, decreased appetite, decreased weight, view of myself, psychomotor slowing, and psychomotor agitation, mood worse in the morning, and distinct quality of mood (Lamers, Milaneschi et al. 2020). The first six of these items are present on the QIDS-SR (items 3, 6, 8, 11, 15, and 16, respectively). There were no substitutable items from the PReDICT scales for the last two symptoms on the NESDA Melancholia dimension. Thus, the current analysis used a Melancholia dimension score range of 0-18, with a higher score being indicative of more severe symptomology.

The Anxious distress dimension was defined by NESDA using three items from the IDS-SR (feeling tense, restless, concentration/worry) and two items from the BAI (fear of awful events and feeling like losing control). The PReDICT scales did not contain items that could be substituted for the BAI items, so we opted to define the Anxious distress dimension in PReDICT using the Psychic Anxiety subscale (items 1-6 and 14, all scored 0-4) of the HAMA (Maier, Buller et al. 1988). We (Brydges et al. in submission) have previously demonstrated the Psychic Anxiety subscale has utility in biological assessments of anxiety. The score range was 0-28, and a higher score indicated more severe symptomology.

### Blood Sampling

The decision to collect blood samples for metabolomics was made after one-third of the PReDICT patients had been enrolled, thus reducing the available sample size from metabolic analyses. Plasma and serum samples were drawn via phlebotomy on the day of randomization without consideration of diet or fasting status and repeated at the week 12 post-treatment visit. Plasma was centrifuged within 10 minutes of blood collection at 4□C. Serum was allowed to clot for 20 minutes and then centrifuged at 4□C. All samples were frozen at -80□C until being thawed for the current analyses.

### Inflammatory Markers

Following Lamers et al. (2020), an *inflammation index* was calculated from plasma levels of CRP and IL-6. Plasma high sensitivity (hs)-CRP concentrations were assayed using the Ultra WR CRP immunoturbidimetric assay from Sekisui Diagnostics (Exton, PA) implemented on the AU480 chemistry analyzer (Beckman Coulter, Brea, CA; detection limit 0.01 mg/L) (Le, Innis-Whitehouse et al. 2000, Mehta, Raison et al. 2013, Raison, Rutherford et al. 2013, Felger, Li et al. 2016, Bekhbat, Chu et al. 2018, Goldsmith, Bekhbat et al. 2020). Concentrations of plasma IL-6 were assessed using high-sensitivity Quantikine enzyme-linked immunosorbent assay (ELISA) kits from R&D systems (Minneapolis, MN; detection limit 0.04 pg/ml) (Raison, Borisov et al. 2009, Raison, Borisov et al. 2010, Musselman, Royster et al. 2013, Smith, Conneely et al. 2014). Assays were performed in duplicate according to the manufacturer’s instructions. Intra- and inter-assay coefficients of variation (CVs) were reliably <10%. To calculate the inflammation index, the CRP and IL-6 measures were each log transformed, standardized, and then averaged, following the procedure described by Lamers et al. (2020).

### Metabolic Profiling

We examined metabolomics associations with MDD using three platforms that provide complementary insights to the metabolism. Metabolomics data focused on primary and polar metabolites using gas chromatography – time of flight mass spectrometry (Fiehn 2016). Briefly, 30 µl of plasma was extracted at -20ºC with 1 mL degassed isopropanol/acetonitrile/water (3/3/2). Extracts were dried down, cleaned from triacylglycerides using acetonitrile/water (1/1), and derivatized with methoxyamine and trimethylsilylation. Samples (0.5 µL) were injected at 250ºC to a 30 m rtx5-SilMS column, ramped from 50-300ºC at 15ºC/min, and analyzed by -70 eV electron ionization at 17 spectra/s. Raw data were deconvoluted and processed using ChromaTOF vs. 4.1 and uploaded to the UC Davis BinBase database (Fiehn, Wohlgemuth et al. 2008) for data curation and compound identification (Lai, Tsugawa et al. 2018). Result data were normalized by SERRF software to correct for drift or batch effects (Fan, Kind et al. 2019).

Bile acids were quantified using targeted metabolomics protocols and profiling protocols established in previous studies (Qiu, Cai et al. 2009, Xie, Wang et al. 2015), and ultra-performance liquid chromatography triple quadrupole mass spectrometry (UPLC-TQMS) (Waters XEVO TQ-S, Milford, USA).

A third metabolite platform with a targeted metabolomics approach used the AbsoluteIDQ® p180 Kit (BIOCRATES Life Science AG, Innsbruck, Austria), with an ultra-performance liquid chromatography (UPLC)/MS/MS system [Acquity UPLC (Waters), TQ-S triple quadrupole MS/MS (Waters)]. This procedure provides measurements of up to 186 endogenous metabolites in quantitative mode (amino acids and biogenic amines) and semi-quantitative mode (acylcarnitines, sphingomyelins, phosphatidylcholines and lysophosphatidylcholines across multiple classes). The AbsoluteIDQ® p180 kit has been fully validated according to European Medicine Agency Guidelines on bioanalytical method validation. Additionally, the kit plates include an automated technical validation to assure the validity of the run and provide verification of the actual performance of the applied quantitative procedure including instrumental analysis. The technical validation of each analyzed kit plate was performed using MetIDQ® software based on results obtained and defined acceptance criteria for blank, zero samples, calibration standards and curves, low/medium/high-level QC samples, and measured signal intensity of internal standards over the plate. De-identified samples were analyzed following the manufacturer’s protocol, with metabolomics labs blinded to the clinical data. The raw metabolomic profiles included 182 metabolite measurements of serum samples. Each assay plate included a set of duplicates obtained by combining approximately 10 µl from the first 76 samples in the study (QC pool duplicates: SPQC) to allow for appropriate inter-plate abundance scaling based specifically on this cohort of samples (n = 24 across all plates). Metabolites with 1) >40% of measurements below the lower limit of detection (LOD) and 2) >30% of SPQC coefficient of variation were excluded from the analysis (n = 160 metabolites passed QC filters). To adjust for the batch effects, a correction factor for each metabolite in a specific plate was obtained by dividing the metabolite’s QC global average by QC average within the plate. Missing values were imputed using each metabolite’s LOD/2 value followed by log2 transformation to obtain a normal distribution of metabolite levels.

### Statistical Analyses

In order to investigate associations between the metabolome, inflammation, and scores on the A/ER, Melancholia and Anxious distress dimensions, partial Spearman rank correlations were conducted between baseline intensity of each compound, the inflammatory index, and the three symptom dimensions after accounting for age, sex, and body mass index (BMI). The resulting correlation coefficients and *p* values were also entered into the ChemRICH tool (Barupal and Fiehn 2017) to test for statistically significant enrichment in sets of structurally similar chemicals using the Kolmogorov-Smirnov statistics. All reported *p* values are uncorrected for multiple comparisons. Additionally, due to the complex relationship between BMI, metabolic alterations and specific atypical energy-related depressive symptoms, all analyses were repeated without accounting for BMI. It is known indeed that risk variants for metabolic pathway alterations (e.g. LEP-R, MC4R) and depression (e.g. NEGR1 [Neuronal Growth Regulator 1]) have a major role in BMI genetic architecture, determining substantial genetic covariance between BMI, metabolic alterations and atypical symptoms (Milaneschi, Lamers et al. 2017, Badini, Coleman et al. 2020, Kappelmann, Arloth et al. 2021, Milaneschi, Lamers et al. 2021). Note that the MDD dimensions are not mutually exclusive “subtypes” of MDD; analyses of all three dimensions use all 158 patients’ scores on each dimension.

We also used Bayes factors (BFs) to investigate the non-associations of inflammation and Melancholia and Anxious distress symptoms (Wagenmakers, 2007). BFs are advantageous over traditional *p* values because they allow for evidence in support of one hypothesis over another to be quantified (including evidence supporting the null hypothesis). Conversely, a *p* value is limited in that a non-significant *p* value cannot distinguish between a true null effect and an effect that is underpowered. Following Jeffreys’ (1961) guidelines, a BF of at least 3 was used as a cut-off for sufficient evidence in favor of the alternative hypothesis (i.e., an association between inflammation and a symptom dimension), and less than 1/3 was considered as sufficient evidence in favor of the null hypothesis. That is, one hypothesis had to be at least three times more likely, given the data, to be considered sufficient evidence in favor of that hypothesis. BFs between 1/3 and 3 were considered as weak evidence, which is indicative of an underpowered effect, and a BF of 1 implies that the null and alternative hypotheses are equally likely, given the data. A scaled beta (1/3, 1/3) distribution was used as the prior distribution.

## RESULTS

### Patient characteristics

Of the 344 patients randomized in PReDICT, 158 had all measures available for analysis at baseline. The demographic and clinical characteristics of these subjects are presented in Table 1.

**Table 1.** Subject Demographic and Clinical Characteristics

| Variable | *M (SD) (N = 158)* |
| --- | --- |
| Age (years) | 39.09 (12.00) |
| Sex (N, % female) | 102 (64.56%) |
| Body mass index | 28.65 (6.47) |
| Race |  |
| White | 62 (39.24%) |
| Native American | 44 (27.85%) |
| Black | 32 (20.25%) |
| Asian | 2 (1.27%) |
| Multiracial | 10 (6.33%) |
| Unknown/Not Reported | 8 (5.06%) |
| Hispanic ethnicity (%) | 57 (36.08%) |
| Immunometabolic dimension score (baseline; max score = 15) | 5.25 (2.57) |
| Melancholia dimension score (baseline; max score = 18) | 5.73 (2.68) |
| Anxious distress dimension score (baseline; max score = 28) | 10.62 (2.75) |

### Associations between Depression Symptoms Dimensions and Inflammation

Table 2 shows the correlations and Bayes Factor (BF) values between the three symptom dimension scores and the inflammation index. IMD symptom dimension scores significantly and positively correlated with inflammation marker levels, but Melancholia and Anxious distress scores did not. Additionally, the BF values for Melancholia and Anxious distress indicated moderate evidence in favor of the null hypothesis (i.e., no association), replicating the results from the NESDA study (Lamers et al 2020). The fact that the correlations between depression dimensions were also quite low (IMD-Anxious distress *r* = .18; IMD-melancholia *r* = .09; Anxious distress-melancholia *r* =.31), highlights the importance of separating these dimensions and implying that these dimensions may be different diseases.

**Table 2.** Correlations and Bayes Factors between Depression Symptom Dimensions and Inflammation Index

| Symptom Dimension | Age and sex accounted for |  |  | Age, sex, and BMI accounted for |  |  |
| --- | --- | --- | --- | --- | --- | --- |
|  | r | p | BF | r | p | BF |
| IMD | 0.22 | .005 | 7.35 | .19 | .019 | 2.11 |
| Melancholia | -0.09 | .280 | 0.20 | 0.03 | .734 | 0.24 |
| Anxious Distress | 0.06 | .444 | 0.23 | 0.10 | .224 | 0.31 |

*Note.* BF refers to Bayes Factor in favor of the alternative hypothesis over the null hypothesis. A BF < 1/3 indicates moderate evidence in favor of the null hypothesis; a BF > 3 indicates moderate evidence in favor of the alternative hypothesis. All *p* values are uncorrected for multiple comparisons. IMD: Immunometabolic depression.

### Biochemical Profile of Immunometabolic Depression

As shown in Figure 1, IMD dimension scores were significantly positively correlated with indoxyl sulfate and indole-3-lactate (tryptophan metabolites generated from specific gut bacteria), and propane-1,3-diol (another microbial product). A short chain free fatty acid, butyric acid (also a microbial product) and the TCA cycle metabolite fumaric acid were positively correlated with IMD scores. Among the acylcarnitines, the short chain C4:1 and C3:1 and the medium chain C10 showed significant inverse correlations with IMD. Several long chain saturated free fatty acids, C16, C17, C18 were inversely correlated to IMD.

**Figure 1.**
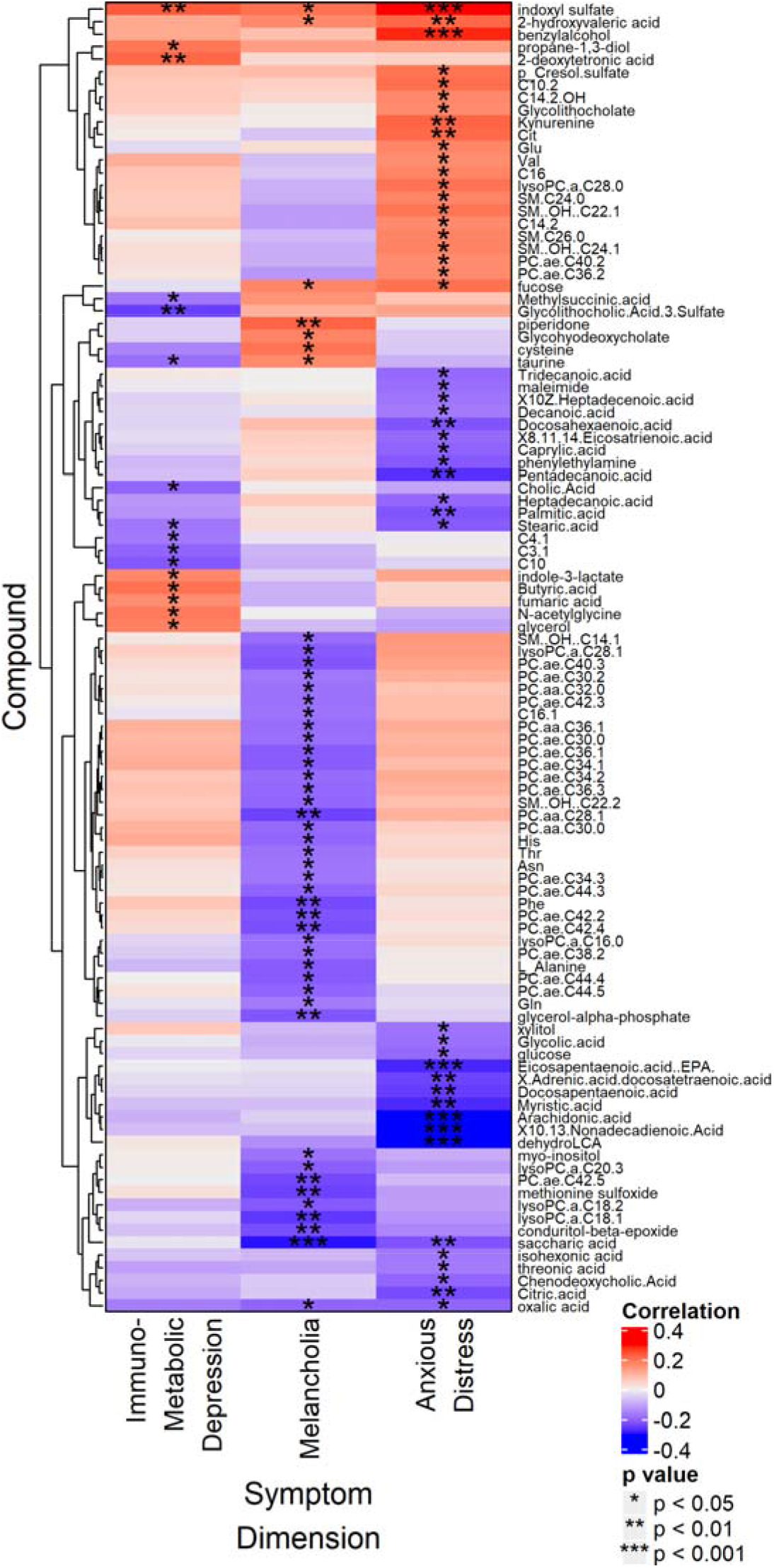
Heat map displaying partial Spearman rank correlations between compound intensity and Immunometabolic, Anxious distress, and Melancholia dimension scores at baseline, after controlling for age, sex, and BMI. Only identified metabolites that correlated significantly with at least one of the three variables are shown. All *p* values are uncorrected for multiple comparisons.

Given the significant correlation between IMD symptom dimension and the inflammatory index, we also compared the metabolomic profiles of the inflammatory index with that of IMD, but few metabolites showed significant correlations with both measures. One compound that was positively correlated with both IMD and the inflammatory index was 2-deoxytetronic acid, a gamma-aminobutyric acid (GABA) breakdown metabolite. One bile acid produced by the gut bacteria, glycolithocholic acid-3-sulfate, was inversely correlated with both measures.

### Biochemical profile of the Melancholia Dimension

The symptom dimension of Melancholia showed significant inverse correlations to several long chain, saturated and unsaturated fatty acyl containing phosphatidylcholines (PCs), and lysoPCs, with or without adjusting for BMI (Figure 1 and supplemental Figures). Among the significantly inversely correlated PCs were a few diacyl ester-linked PCs, including PCaaC28:1, -C30:0, -C32:0, -C36:1, -C38:3. However, the majority of the PCs that showed inverse correlation to melancholia were presumably the ether-linked PCs such as PCaeC30:0, -C30:2, -C34:1, -C34:2, -C34:3, - C36:1, -C36:2, -C36:3, -C38:2, -C40:2, -C40:3, -C42:2, -C42:3, C42:4, -C42:5, -C44:3, - C44:4, -C44:5, -C44:6. Among the ester lysoPCs, -C16:0, -C18:1,-C18:2, -C20:3, - C28:1 were inversely correlated to severity of Melancholia.

Besides the lipids, several amino acids, including asparagine, glutamine, histidine, phenylalanine, threonine, and alanine, were significantly negatively correlated to the Melancholia dimension score. Saccharic acid, a derivative of glucose, was also significantly inversely correlated with Melancholia severity. In contrast, the gut microbiome-derived tryptophan metabolite indoxyl sulfate was significantly positively correlated to severity of Melancholia, as was 2-hydroxyvaleric acid, a highly hydrophobic, short chain hydroxyl fatty acid.

### Biochemical profile of the Anxious Distress Dimension

The Anxious distress dimension was significantly negatively correlated with several medium and long chain free fatty acids (FFAs), both saturated and unsaturated, including polyunsaturated fatty acids (Figure 1). Among the saturated FFAs were the medium chain C8:0 and C10:0, and the long chain C13:0, C14:0, C15:0, C16:0, C17:0, and C18:0 with both odd-chain FAs and even-chain FAs showing significant correlations to the symptom severity. Among the polyunsaturated fatty acids there were the omega-3 FFAs like Docosahexaenoic acid (DHA), Docosapentaenoic acid (DPA), Eicosapentaenoic acid (EPA), and several omega-6 FFAs as well, e.g., arachidonic acid (AA), C19:2(cis_10,13), C20:3(cis_8,11,14) and C22:4(cis_7,10,13,16). However, no significant associations of this dimension were detected with the ratios of omega3/omega-6 FFAs. In contrast, several amino acid levels, including citrulline, glutamate, valine, and kynurenine, were all significantly positively correlated with this dimension. Gut-microbiome-derived secondary bile acids like glycolithocholic acid and its sulfate also showed positive correlations with severity of anxiety distress, whereas the primary bile acid, CDCA showed opposite relationship. Other gut-microbiome related metabolites that were significantly positively correlated to anxiety severity were indoxyl sulfate, p-cresol sulfate, benzyl alcohol, and 2-hydroxyvaleric acid. Finally, several medium and long chain acylcarnitines (C10:2, C14:2, C14:2OH, C16) as well as a number of hydroxy-sphingolipids were positively correlated with Anxious distress scores.

### Comparison across MDD dimensions and inflammation

Indoxyl sulfate was the only metabolite that showed a significant positive correlation to all three symptom dimensions, with or without controlling for BMI (Figure 1). Other metabolites significantly correlated in the same direction with both Melancholia and Anxious distress were: 1) 2-hydroxyvaleric acid (positive correlations), 2) fucose, a monosaccharide (positive correlations), 3) oxalic acid (negative correlations), and 4) saccharic acid, a hydrophobic glucuronic acid derivative (negative correlations). The only compound significantly correlated with both IMD and Melancholia was taurine, though the correlations were in opposite directions: Melancholia (positive) and IMD (negative).

This low level of overlap in biochemistry across these clinical dimensions was further supported by the ChemRICH enrichment analyses (Figure 2). The heatmap shows that no chemical clusters were common to any of the three symptom dimensions with the exception of sphingolipids for Melancholia and Anxious distress, but in that case the associations were opposite directions.

**Figure 2.**
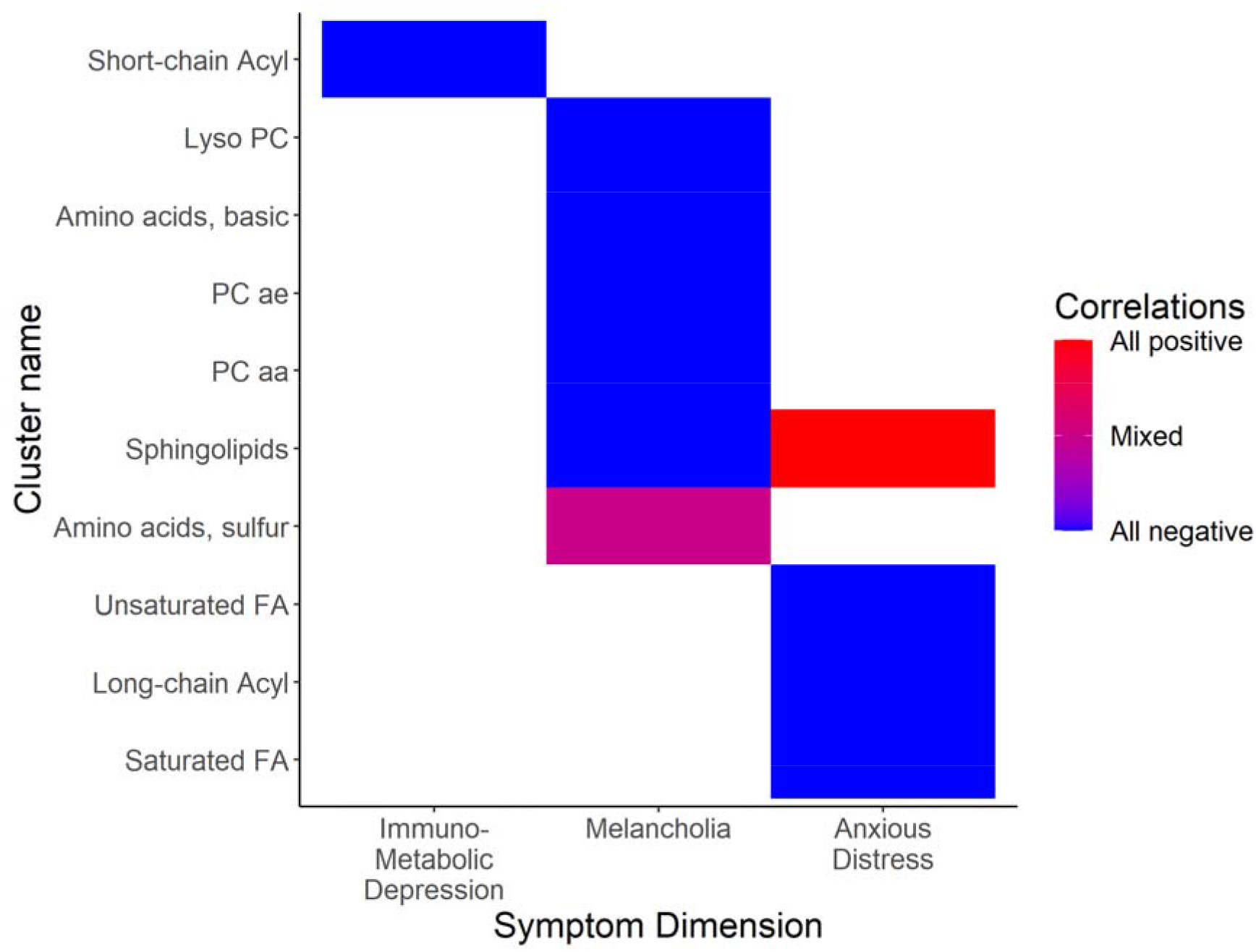
ChemRICH enrichment results for Immunometabolic, Melancholia, and Anxious distress dimensions after controlling for age, sex, and BMI. The color of each cell indicates the direction of the correlations in the chemical cluster (blue = all compounds negatively correlate with phenotype; red = all compounds positively correlate with phenotype; pink/purple = correlations in both directions). *Note*. PC aa: diacyl ester-linked phosphatidylcholines. PC ae: ether-linked phosphatidylcholines. FA: fatty acids.

## Discussion

The results of the analyses reported here identify two important findings for understanding the heterogeneity of MDD. First, we replicated the NESDA study findings of a specific association of the inflammatory index (defined by plasma CRP and IL-6 concentrations) with the clinical dimension of IMD, and the lack of association of the inflammatory index with the Melancholia and Anxious distress dimensions (Lamers et al., 2020). This replication provides further support for the pathogenic impact of elevated states of inflammation for certain MDD patients, particularly those who report very low energy despite increases in appetite and sleep as part of their major depressive episode. Second, we identified specific, minimally overlapping metabolomic associations of the three MDD clinical dimensions. These biochemical differences are likely to contribute to the biological processes that underlie the variability in clinical presentations of MDD. Taken together, this and others’ work (Ahmed, MahmoudianDehkordi et al 2020, Liu, Yieh et al 2016, Alshehri, Mook-Kanamori et al 2021, Kunugi, Hori et al 2015) demonstrates the utility of using biochemistry for dissecting the heterogeneity of MDD.

The IMD dimension was positively associated with several gut metabolites (i.e., high metabolite levels were associated with more severe symptomology), including indoles that are derived from tryptophan metabolism. Tryptophan is an essential amino acid that is metabolized in the gastrointestinal tract via the indole, serotonin, and kynurenine metabolic pathways, which have been associated with a range of health and neuropsychiatric pathologies including inflammatory diseases, metabolic syndrome, depression, and anxiety disorders (Agus, Planchais et al. 2018, Gao, Xu et al. 2018). For example, indoxyl sulfate is derived from tryptophanase-producing bacteria and has previously been found to be associated with depression (Philippe, Szabo de Edelenyi et al. 2021). Additionally, indole-3-lactic acid and other gut microbiome-linked metabolites such as propane-1,3-diol, which is produced from glycerol by *Clostridium diolis* bacteria and *Enterobacteriaceae*, were also positively associated.

Of interest was the positive correlation between 2-deoxytetronic acid, a metabolite formed through a GABA degradation pathway, and severity in the IMD dimension. 2-deoxytetronic acid is a marker of succinic semialdehyde dehydrogenase deficiency (Shinka, Inoue et al. 2002) that is associated with a rare autosomal recessive disorder and may be informative about Alzheimer disease (Mousavi, Jonsson et al. 2014).

IMD severity was positively correlated with concentrations of the TCA cycle metabolite fumaric acid, and negatively correlated with several acylcarnitines, including the short chain C4:1 and C3:1 and the medium chain C10. A striking observation in this study was the significant association of free fatty acids and lipids, particularly those implicated in membrane structure and function, with the dimensions of anxiety distress and melancholia, respectively, but not with the IMD dimension.

Patients with greater Melancholia dimension severity were characterized by significantly low baseline levels of phosphatidylcholines such as several diacyl PCs, lysoPCs but mostly the ether-linked phosphatidylcholines. Almost 80% of the PCs that were significantly associated with severity of Melancholia were ether-linked. A major class of ether-linked phospholipids are plasmalogens that are ubiquitous in mammalian cell membranes and plasma lipoproteins. They are enriched in the myelin sheath and are key components of “lipid rafts” (Di Paolo and Kim 2011, Dean and Lodhi 2018, Lefevre-Arbogast, Hejblum et al. 2021). They are relevant for these specific membrane microdomains, which act as dynamic platforms for several cellular processes, including signaling pathways, molecular trafficking and protein interactions (Di Paolo and Kim 2011) and play critical role in neurotransmission and synaptic plasticity (Sebastiao, Colino-Oliveira et al. 2013).

Ether-linked PCs also act as potent anti-oxidants in cellular membranes by preventing oxidative degradation of polyunsaturated fatty acid containing phospholipids (Liu, Yieh et al 2016, Dorninger, Forss-Petter et al 2020, Dean and Lodhi 2018). The reduced levels of ether-linked PCs in Melancholia may thus be indicative of an increased oxidative stress, related to cellular membrane structure, in these patients rather than enzymatic inactivity at the phospholipase A2 (PLA2) level. PLA2 participates in fatty acyl chain remodeling of phospholipids through fatty acyl hydrolysis, releasing lysoPCs and FFAs. In the more severe Melancholia patients not only the PCs were low but the end-products of PLA2 activity, lysoPCs and FFAs were either significantly or trended to be low (Figure 1) which suggests changes in PLA2 activity is probably not the mechanism here. We considered the possibility of increased activity of lecithin cholesterol acyltransferase (LCAT) to explain these results but rejected this hypothesis because LCAT uses PCs to esterify free cholesterol on high density lipoproteins (HDL) thereby releasing lysoPCs, and this process is not selective for ether-lipids, as our data demonstrates. Thus, the reduced levels of both PC and lysoPC and low or unchanged masses of FFAs appear more likely due to defects in PC biosynthesis, for example, in the *de novo* synthesis of phospholipids via the Kennedy pathway and/or the biosynthesis of plasmalogen precursor. This explanation better fits the observed low levels of both PC and lysoPC since PC serves as the precursor of lysoPC production. This hypothesis of a defect in PC biosynthesis will need to be tested with measurement of liver function including hepatic peroxisomal activity and/or the HDL levels. Whatever the explanation, plasmalogens are thought to play a significant role in Alzheimer’s disease (Su, Wang et al. 2019) and other genetic neurological and metabolic diseases. However, whether they play a causal role or represent a consequence of the underlying pathophysiology is yet to be ascertained (Paul, Lancaster et al. 2019).

In contrast to the findings for the Melancholia dimension, higher levels of PCs with both the diacyl-ester linked and the ether-linked fatty acyl chains were associated with greater scores on the Anxious distress dimension. Although this association did not reach statistical significance, overall they were positively correlated to more severe symptoms of anxiety. Significantly low levels of the serum free fatty acid levels, both saturated and unsaturated, were also be observed in these patients. This may suggest a possible disruption in the activity of phospholipase A2 function. Phospholipases A2 (PLA2s) are enzymes that cleave fatty acid in position two of phospholipids, hydrolyzing the bond between the second fatty acid “tail” and the glycerol molecule (Figure 3), thus releasing a free fatty acid and a lysophosphatidic acid. In particular, PLA2 has been shown to release arachidonic acid that, upon downstream modification by cyclooxygenases or lipoxygenases, is modified into active compounds called eicosanoids (Figure 3). Eicosanoids include prostaglandins and leukotrienes, which can function as both anti-inflammatory and pro-inflammatory mediators by regulating the immune response (Dennis 1994). We found a highly significant inverse correlation between Anxious distress scores and levels of arachidonic acid, suggesting a possible role of inflammatory mediators for this dimension. However, we did not detect significant correlations between the inflammatory index and the Anxious distress dimension score.

**Figure 3.**
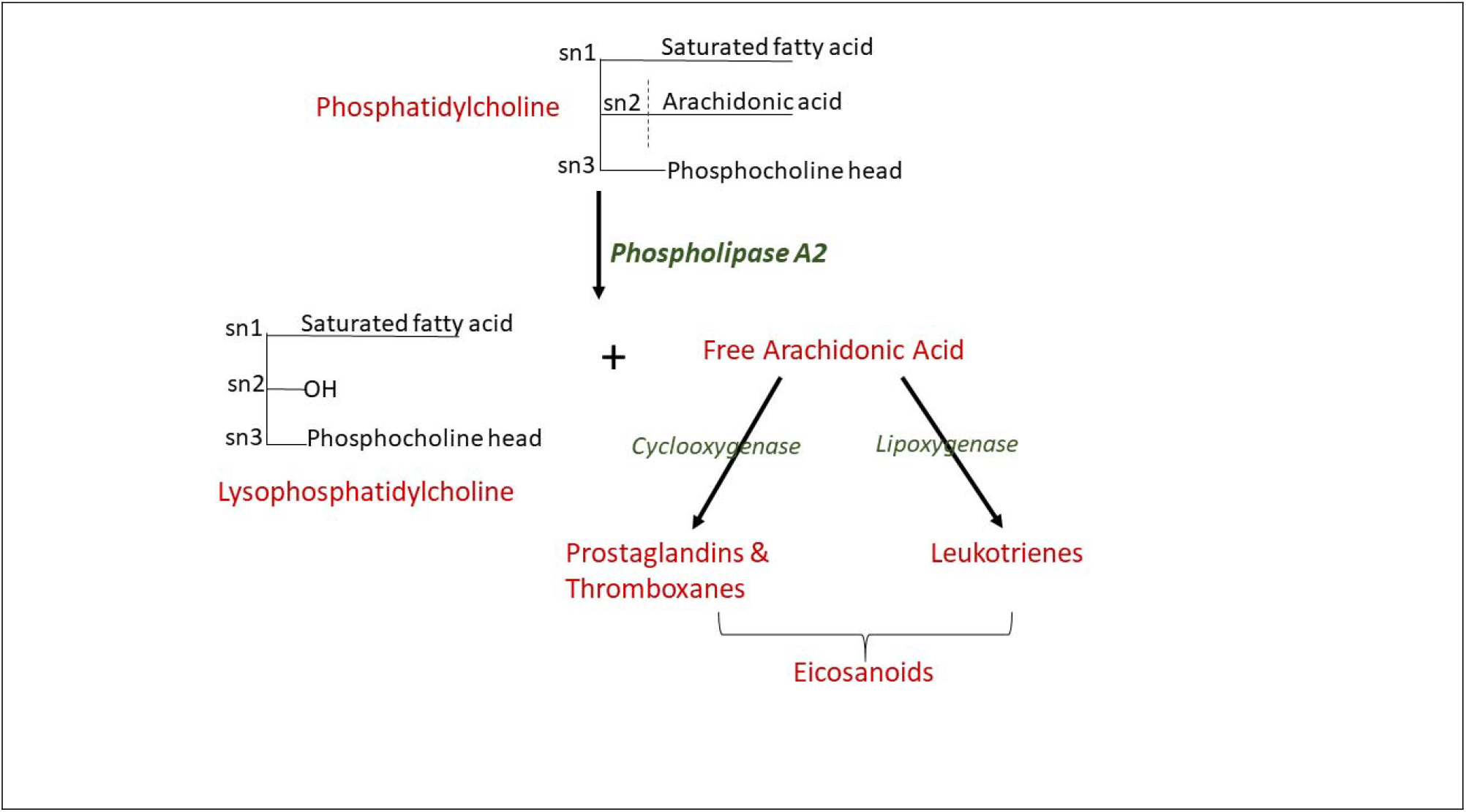
Pathway showing phosphatidylcholine metabolism by phospholipase A2 to produce lysophosphatidylcholine and a free fatty acid, particularly Arachidonic Acid which goes on to produce the inflammation-mediating eicosanoids.

Additionally, several other polyunsaturated free fatty acids were also significantly inversely correlated to severity of anxiety, such as EPA, DHA, and docosapentaenoic acid. DHA, followed by its precursor EPA, are the most common omega-3 fatty acids in human body (Schmitz and Ecker 2008, Parekh, Smeeth et al. 2017). DHA alone comprises 15%–20% of human brain lipid and lower concentrations in this group has been shown to have strong association to depression due to dysregulation of inflammatory responses, decreased antioxidant capacity and disrupted neurotransmission (Parekh, Smeeth et al. 2017). Several previous reports have indicated that patients with MDD have a lower level of the PUFAs such as EPA and DHA in their peripheral tissues (plasma, serum and red blood cells) than control subjects, and furthermore, the severity of depressive symptoms is correlated with lower omega-3 PUFAs (Edwards, Peet et al. 1998). Here we show that among the MDD patients, specifically the Anxious distress dimension, there were significant inverse correlations of EPA, DHA with increased severity of the symptoms. PUFAs are important components of phospholipids and cholesterol esters of the neuronal cell membrane, especially of dendritic and synaptic membranes and constitute principal regulating factors of neurogenesis, cell survival and neurotransmission. Higher circulating *n*-3 PUFAs are known to be associated with a lower risk for metabolic syndrome. This syndrome refers to a cluster of three or more components of cardiometabolic risk factors, including central obesity, hypertension, dyslipidemia and impaired glucose tolerance. Quality of dietary fat is known to play significant role in the development of metabolic syndrome. Both omega-3 and omega-6 PUFAs are inversely associated with more severe anxiety distress in our study which may suggest presence of metabolic syndrome in these patients, partly secondary to poor dietary sources. However genetic or hormonal factors may also play a role. Additionally, the PUFAs being low may result from increased lipid peroxidation from reactive oxygen species due to increase in oxidative stress.

Besides the phospholipids and the FFAs, several medium and long chain acylcarnitines such as C10:2, C14:2OH, C16 and C14:2 were also higher in more severely anxious patients, likely indicating disrupted beta-oxidation in the mitochondrial energy metabolism in these patients. We have previously reported a decrease in several medium and long chain acylcarnitines after SSRI treatment within the clinical subtype of anxiety in a separate MDD cohort (Ahmed, MahmoudianDehkordi et al. 2020, MahmoudianDehkordi, Ahmed et al. 2021), suggesting that the SSRI, Citalopram/Escitalopram treatment may restore beta-oxidation related mitochondrial dysfunction in clinically anxious MDD patients.

Among other molecules of interest in Anxious distress dimension were the significant positive correlations of several gut-bacteria derived metabolites such as indoxyl sulfate, p-cresol sulfate, 2-hydroxyvaleric acid, benzyl-alcohol and the toxic bile acid glycolithocholate with increased symptom severity, suggesting gut-dysbiosis in these patients.

Among the amino acids, kynurenine and citrulline showed positive correlations to severity of anxious distress. Our previous work with our colleagues at Mayo Clinic has shown that kynurenine, a tryptophan metabolite, was strongly associated with depressive symptoms in the Mayo-PGRN cohort of 290 MDD outpatients (Liu, Ray et al. 2018). It has been observed that inflammatory cytokines can increase expression of the enzyme indoleamine 2,3-dioxygenase, thus activating the kynurenine branch of tryptophan metabolism resulting in depressive-like behavior in mouse model (O’Connor, Andre et al. 2009).

Citrulline is an intermediate of the urea cycle and previous studies have observed significantly low levels of arginine and citrulline in MDD patients compared to healthy controls (Hess, Baker et al. 2017). We have shown increase in these urea cycle intermediates in MDD patients in response to exposure to the SSRI escitalopram/citalopram (MahmoudianDehkordi, Ahmed et al. 2021) which may be indicative of alterations in urea cycle, removal of nitrogen waste and nitric acid production in these patients.

There are some limitations to these analyses. Although the sample size included over 150 treatment naïve MDD adults, we did not have sufficient power to meaningfully apply corrections for multiple comparisons. We could not control for diet and the samples were collected without regard to fasting status, which may have particularly impacted some of the lipids reported. Another limitation is that the Biocrates p180 kit coupled with the mass-spectrometer platform used cannot discern lipid molecules with very small mass differences (i.e., less than 0.05 Da) and therefore it is possible that the SM(OH)s might be long-chain SMs instead (Li, Alam et al. 2021). Similarly, the platform also has limited resolution to differentiate between long chain lysophosphatidylcholines and phosphatidylcholines.

In conclusion, this study indicates that a multifaceted disruption in the delicate balance between the gut microbiota, dietary lipids and host lipid metabolism triad relationship may be a cause of distinct symptom dimensions of MDD. Considering that all patients contributed to all metabolome-symptom dimension comparisons, the distinctness of the metabolomic profile for each dimension is noteworthy.

## Data Availability

Access to Data will be made available upon requests.

## Acknowledgements

We acknowledge the assistance of Ms. Lisa Howerton (Duke). This work was funded by grant support to Dr. Rima Kaddurah-Daouk (PI) through NIH grants R01MH108348, R01AG046171 and U01AG061359. Dr. Boadie Dunlop has support from NIH grants P50-MH077083 (PI Mayberg), R01-MH080880 (PI Craighead), UL1-RR025008 (PI Stevens), M01-RR0039 (PI Stevens) and the Fuqua Family Foundations. The PReDICT study was supported by NIH Grants numbered: P50 MH077083 and RO1 MH080880. Dr. Penninx has received research funding (unrelated to the work reported here) from Jansen Research and Boehringer Ingelheim. Drs. Gabi Kastenmüller and Matthias Arnold have support from NIH grants U01AG061359 (PI Kaddurah-Daouk), 1RF1AG059093 (PI Kaddurah-Daouk), 1RF1AG058942 (PI Kaddurah-Daouk), and 1U19AG063744 (PI Kaddurah-Daouk). Dr. Bhattacharyya had support for this work from the NIH grant R01MH108348 (PI Kaddurah-Daouk and Dunlop).

